# Pre-pandemic Cognitive Function and COVID-19 Vaccine Hesitancy: Cohort Study

**DOI:** 10.1101/2021.03.16.21253634

**Authors:** G. David Batty, Ian J. Deary, Chloe Fawns-Ritchie, Catharine R. Gale, Drew Altschul

## Abstract

**Background:** Whereas several predictors of COVID-19 vaccine hesitancy have been examined, the role of cognitive function following the widely publicised development of an inoculation is unknown.

**Objective:** To test the association between scores from an array of cognitive function tests and self-reported vaccine hesitancy after the announcement of the successful testing of the Oxford University/AstraZeneca vaccine.

**Design, Setting, and Participants:** We used individual-level data from a pandemic-focused study (COVID Survey), a prospective cohort study nested within Understanding Society (Main Survey). In the week immediately following the announcement of successful testing of the first efficacious inoculation (November/December 2020), data on vaccine intentionality were collected in 11740 individuals (6702 women) aged 16-95. Pre-pandemic scores on general cognitive function, ascertained from a battery of six tests, were captured in 2011/12 wave of the Main Survey.

**Measurements:** Self-reported intention to take up a vaccination for COVID-19. To summarise our results, we computed odds ratios with accompanying 95% confidence intervals for general cognitive function adjusted for selected covariates.

**Results:** Of the study sample, 17.2% (N=1842) indicated they were hesitant about having the vaccine. After adjustment for age, sex, and ethnicity, study members with a lower baseline cognition score were markedly more likely to be vaccine hesitant (odds ratio per standard deviation lower score in cognition; 95% confidence interval: 1.76; 1.62, 1.90). Adjustment for mental and physical health plus household shielding status had no impact on these results, whereas controlling for educational attainment led to partial attenuation but the probability of hesitancy was still elevated (1.52; 1.37, 1.67). There was a linear association for vaccine hesitancy across the full range of cognition scores (p for trend: p<0.0001).

**Limitations:** Our outcome was based on intention rather than behaviour.

**Conclusions:** Erroneous social media reports might have complicated personal decision-making, leading to people with lower cognitive ability test scores being vaccine-hesitant. With people with lower cognition also experiencing higher rates of COVID-19 in studies conducted prior to vaccine distribution, these new findings are suggestive of a potential additional disease burden.

## Introduction

Cognitive function – also known as mental ability or intelligence – refers to psychological functions that involve the storage, selection, manipulation, and organisation of information, and the planning of actions.^1,2^ Assessed using standard tests, there is marked inter-person variation in how rapidly and precisely people carry out these mental tasks.^1,2^ Health protection and health care can also be regarded as a complex set of assignments that require assimilation of knowledge, decision-making, and planning. It has been posited that people with higher cognitive function manage preventative behaviours and treatment more effectively,^3^ and there is growing evidence that this is the case.

In well-characterised cohort studies, relative to their lower-performing counterparts, people with higher ability are more likely to have a healthy diet,^4^ choose dietary supplements,^5^ and be physically active.^4^ Those who score better on cognitive tests also have a lower probability of smoking cigarettes,^6,7^ drinking harmful levels of alcohol,^8^ and having associated problems.^9^ Cessation rates are also elevated in smokers with higher mental ability.^10^ Further, in individuals with a greater risk of a first cardiovascular disease event,^11^ in those with a higher probability of re-infarction,^12^ and in patients with respiratory disease,^13^ improved compliance with known efficacious drug therapies is apparent with higher ability scores. Similarly, in people with an elevated risk of colorectal cancer, rates of participation in a free screening programme were elevated in persons with better performance on tests of cognition.^14^

These observations provide circumstantial evidence for a link between cognitive ability and another health-protecting behaviour, vaccine uptake. Vaccination is central to controlling the present pandemic, with success reliant on a sufficiently high uptake to achieve herd immunity.^15^ In the only empirical investigation of which we are aware, investigators administered a very brief measure of analytical reasoning to people in two small cross-sectional studies from the UK (N=2025) and Ireland (N=1041).^16^ Relative to the group who indicated they would be likely to accept a COVID-19 inoculation if one became available, somewhat lower cognition scores were apparent in study members indicating vaccine reticence.^16^ These data were collected in March/April 2020 when no vaccine was available. Around 8 months later, following periodic statements to the media of its on-going development, successful testing of the Oxford University/AstraZeneca vaccine, the first known efficacious inoculation against COVID-19, was announced.^17^ Time-series analyses across multiple countries potentially implicates this declaration and those for other vaccines in having a positive impact upon intentionality.^18^ Accordingly, for the first time to our knowledge, we investigated the link between cognitive function and COVID-19 vaccine hesitancy in a large UK general population-based sample in which data collection took place following the announcement of an efficacious vaccine.

## Methods

Understanding Society, also known as the UK Household Longitudinal Study, is a nationally-representative, on-going, open, cohort study (hereinafter, the ‘Main Survey’). Based on a clustered-stratified probability sample of households, participants have been interviewed annually since 2009.^19^ Households who had participated in at least one of the two most recent waves of data collection (wave 8, 2016-18; wave 9, 2017-19) comprised the target sample for a pandemic-focused study initiated in April 2020 (hereinafter, the ‘COVID Survey’).^20,21^ The derivation of the present analytical sample from the Main and COVID surveys, including the whereabouts of relevant data, is depicted in figure 1. The University of Essex Ethics Committee gave approval for data collection in the COVID-orientated surveys (ETH1920-1271); no further ethical permissions were required for the present analyses of anonymised data.

**Figure 1.**
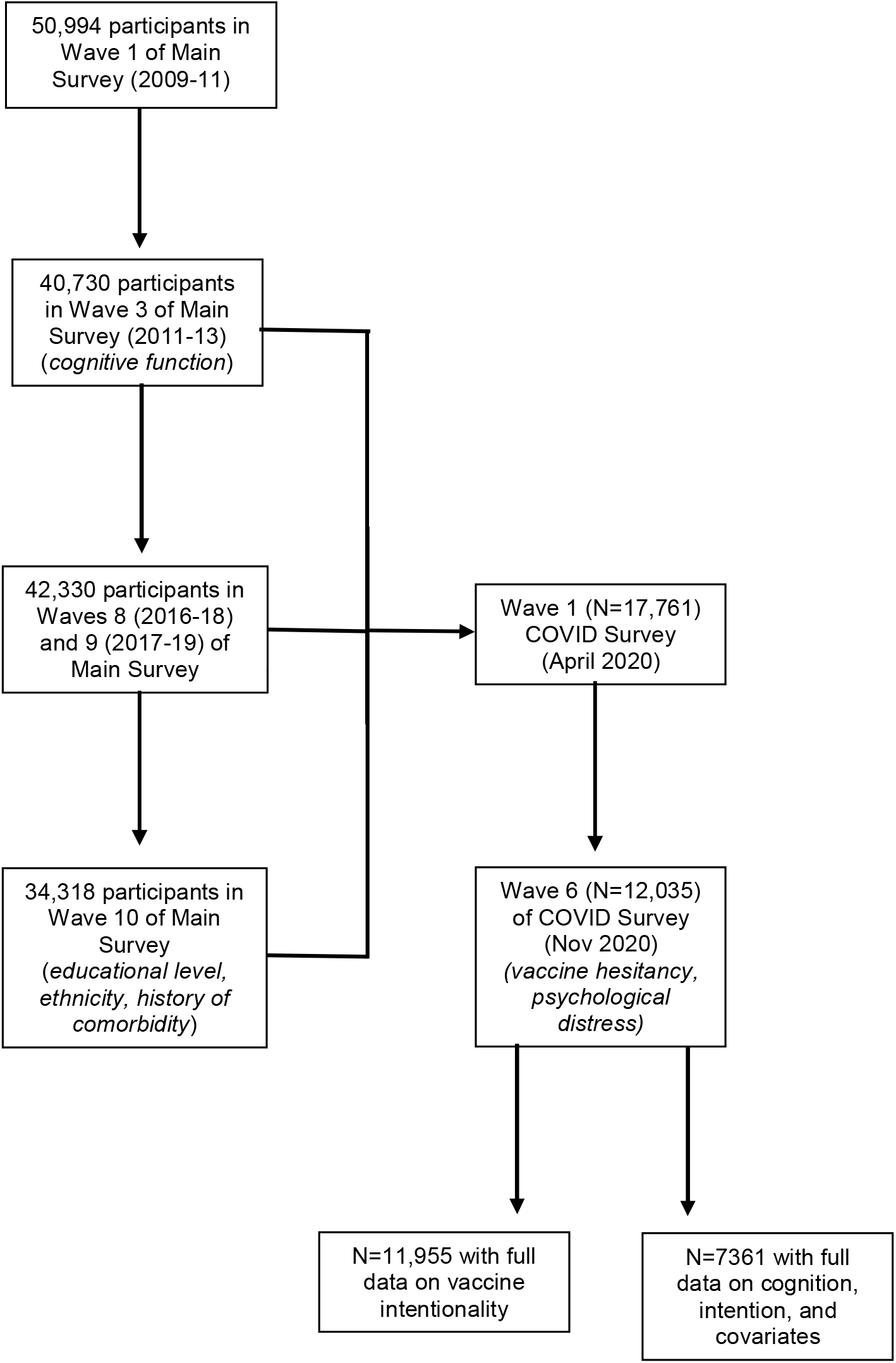
Flow of cohort members into the analytical sample: Main Survey and COVID Survey in Understanding Society.

The COVID Surveys took place monthly/bimonthly between April (wave 1) and November 2020 (wave 6), with questions on vaccine intention first administered in the November wave when study members were aged 16-95 (mean 53).^21^ Data collection in wave 6 (starting 24th November) commenced the day immediately following the announcement of the efficacy of the Oxford University/AstraZeneca vaccine^17^ and continued for one week, comprising a total of 12,035 individuals of 19,294 invitations issued (response proportion, 62%).^21^

### Assessment of cognitive function

In the third wave of data collection in the Main Survey (2011-2013), six cognitive function tests were administered following piloting.^22,23^ Representing a range of cognitive skills, these tests have been repeatedly deployed in large-scale, population-based studies.^24-28^ Verbal declarative memory was assessed using both *immediate word recall* and *delayed word recall* tasks. Respondents listened to a list of ten words delivered by a computer; they were then asked to immediately recall the words and, again, at a later stage in the interview without having heard the words again. The number of correct responses was recorded on each occasion. For *semantic verbal fluency*, respondents named as many animals as they could in one minute; the final score was based on the number of unique correct responses. Using components of screening instruments for *cognitive impairment* including the Mini Mental State Examination^29^ and the Cambridge Cognitive Examination,^30^ respondents were asked to subtract 7 from 100 and then subtract 7 from their answer on four more occasions. The number of correct responses of a maximum of five was recorded. *Fluid reasoning* was assessed using a number sequence in which the respondent populated the gap(s) in a logical series. Respondents were initially presented with simple examples to test their understanding; those who seemed confused or unable to understand test requirements after the relaying of two examples took no further part in the test. Remaining study members were administered two sets of three number sequences, with the difficulty of the second set determined by their performance on the first. A score was derived which accounts for the difficulty of the items. For *numerical reasoning skills*, individuals were given three numerical problems to solve and, depending on their responses, were then administered a further one (simpler) or two (more difficult) problems. The total number of correct responses was recorded.

### Assessment of covariates

Covariates were self-reported and included age; sex (both wave 10, Main Survey); ethnicity (wave 10, Main Survey; denoted as white or non-white); highest education level (wave 10, Main Survey; categorised as degree & other higher degree, A’ level or equivalent [Advanced Placement in the US], GCSE or equivalent [Grade 10 in the USA], other qualification, and none); and National Health Service-recommended shielding status for any household member (waves 1-5, COVID Surveys; denoted by yes/no). A history of physical morbidities was also captured (wave 10, Main Survey) and based on any mention of a cardiometabolic condition (congestive heart failure, coronary heart disease, angina, heart attack or infarction, stroke, diabetes, and/or hypertension); respiratory disease (respiratory disease comprised bronchitis, emphysema, chronic obstructive pulmonary disease, and/or asthma); or cancer of any type. Current psychological distress (wave 6, COVID Survey) was ascertained using the administration of the 12-item version of the General Health Questionnaire. Validated against standardised psychiatric interviews,^31,32^ this is a widely used measure of distress in population-based studies. Consistent with published analyses,^33-35^ we used a score of ≥3 to denote psychological distress.

### Assessment of vaccine intentionality

At wave 6 in the COVID Survey, study members were asked “Imagine that a vaccine against COVID-19 was available for anyone who wanted it. How likely or unlikely would you be to take the vaccine?”. Possible responses were “Very likely”, “Likely”, “Unlikely” and “Very unlikely”. The latter two categories were combined to denote vaccine hesitancy.

### Statistical analyses

It is well-replicated that performance on tests of cognitive abilities are positively inter-related, whereby people who score highly on one test of cognition tend to score well on another.^2^ This has led to the use of the term ‘general cognitive ability’, usually known as ‘g’. Accordingly, using scores from the six tests of cognitive function we generated a single general cognitive function variable. Computed using principal components analysis, the first unrotated component of the six cognitive tests was used as a single measure of cognitive function (variance explained: 42%; loadings: immediate recall 0.74, delayed recall 0.72, verbal fluency 0.59, serial sevens 0.49, number series 0.64, numerical problem solving 0.66). To summarise the relation between cognition and vaccine hesitancy, we used logistic regression to compute odds ratios with accompanying 95% confidence intervals. In these analyses we calculated effect estimates for tertiles of cognitive function scores and those for a unit (standard deviation) disadvantage in score. The most basic analyses were adjusted for age, sex, and ethnicity. Retaining these covariates, we then explored the impact of separately controlling for existing somatic medical conditions, mental health, education, and shielding status.

### Patient involvement

The present analyses are based on existing data of a typically healthy population. To our knowledge, no patients were involved in setting the present research question or the outcome measures, nor were they involved in developing plans for recruitment, design, or implementation of the study. Additionally, no patients were asked to advise on interpretation or writing up of results. Results from Understanding Society are routinely disseminated to study participants via the study website.

## Results

In a sample of 11,955 individuals who responded in full to the enquiry regarding COVID-19 vaccine intentionality, 17.2% (N=1842) indicated that they were hesitant (figure 1). In table 1 we show unadjusted study member characteristics according to vaccine intention. Relative to the group who indicated a willingness to have the vaccine, those who were hesitant were more likely to be young, female, from an ethnic minority background, and be less well educated. The hesitant were also less likely to carry an array of existing somatic morbidities and be shielding or live with someone who was. The prevalence of psychological distress was somewhat higher in the vaccine hesitant.

**Table 1.**
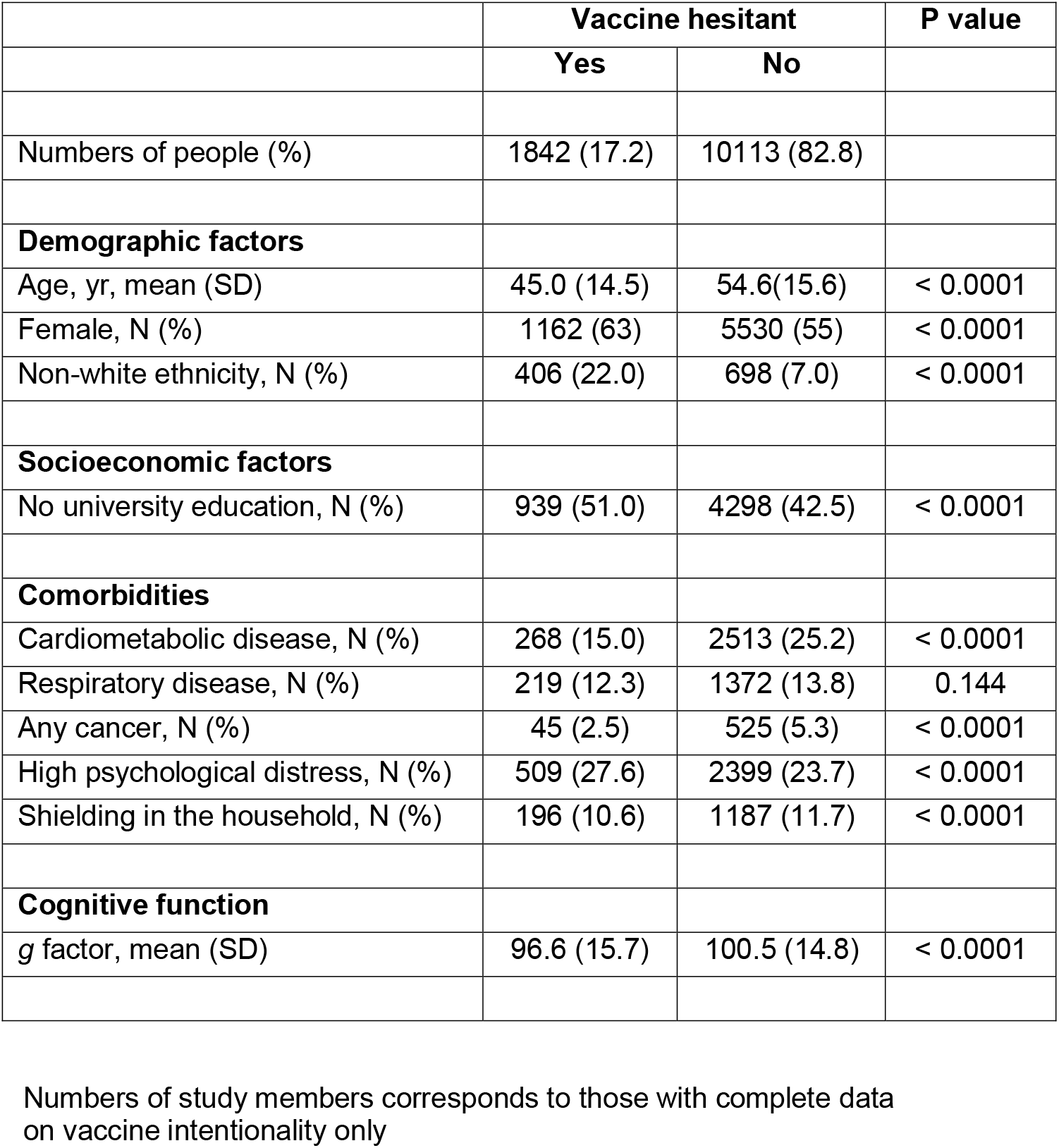
Study member characteristics according to COVID-19 vaccine hesitancy in Understanding Society.

|  | Vaccine hesitant |  | P value |
| --- | --- | --- | --- |
|  | Yes | No |  |
| Numbers of people (%) | 1842 (17.2) | 10113 (82.8) |  |
| **Demographic factors** |  |  |  |
| Age, yr, mean (SD) | 45.0 (14.5) | 54.6(15.6) | < 0.0001 |
| Female, N (%) | 1162 (63) | 5530 (55) | < 0.0001 |
| Non-white ethnicity, N (%) | 406 (22.0) | 698 (7.0) | < 0.0001 |
| **Socioeconomic factors** |  |  |  |
| No university education, N (%) | 939 (51.0) | 4298 (42.5) | < 0.0001 |
| **Comorbidities** |  |  |  |
| Cardiometabolic disease, N (%) | 268 (15.0) | 2513 (25.2) | < 0.0001 |
| Respiratory disease, N (%) | 219 (12.3) | 1372 (13.8) | 0.144 |
| Any cancer, N (%) | 45 (2.5) | 525 (5.3) | < 0.0001 |
| High psychological distress, N (%) | 509 (27.6) | 2399 (23.7) | < 0.0001 |
| Shielding in the household, N (%) | 196 (10.6) | 1187 (11.7) | < 0.0001 |
| **Cognitive function** |  |  |  |
| g factor, mean (SD) | 96.6 (15.7) | 100.5 (14.8) | < 0.0001 |

Numbers of study members corresponds to those with complete data on vaccine intentionality only

There were also differences in cognitive function between the vaccine groups, whereby the vaccine hesitant study members had lower general ability scores (difference in mean score 3.9; p-value for difference: <0.0001). We investigated these differentials in table 2 where we present the results of regression analyses incorporating potential explanatory variables in an analytical sample of 7361 people with full data for the variables depicted (figure 1). In age-, sex- and ethnicity-adjusted analyses, a one standard deviation lower score in general cognitive ability was associated with a 76% greater risk of being vaccine hesitant (odds ratio; 95% confidence interval: 1.76; 1.62, 1.90). While separate adjustments for somatic comorbidity, psychological distress, and shielding had no impact on this relationship, adjustment for education led to some attenuation (1.52; 1.37, 1.67) – the Kendall rank correlation between cognition and educational attainment was 0.27 (p<0.0001). Simultaneous adjustment for all covariates had no greater attenuating effect than the adjustment for education.

**Table 2.** Odds ratios (95% CI) for the relation of general cognitive function with COVID-19 vaccine hesitancy in Understanding Society.

|  | **Number**<br>**hesitant /**<br>**Total at**<br>**risk** | **Age, sex, &**<br>**ethnicity** | **Age, sex,**<br>**ethnicity, &**<br>**somatic**<br>**comorbidity** | **Age, sex,**<br>**ethnicity, &**<br>**psychological**<br>**distress** | **Age, sex,**<br>**ethnicity, &**<br>**shielding** | **Age, sex,**<br>**ethnicity, &**<br>**education** | **All covariates** |
| --- | --- | --- | --- | --- | --- | --- | --- |
| Tertile 3 (high) | 236 / 2048 | 1 (ref) | 1 (ref) | 1 (ref) | 1 (ref) | 1 (ref) | 1 (ref) |
| Tertile 2 | 352 / 2566 | 1.28 (1.07, 1.54) | 1.29 (1.08, 1.55) | 1.28 (1.07, 1.54) | 1.29 (1.08, 1.55) | 1.17 (0.98, 1.41) | 1.18 (0.99, 1.42) |
| Tertile 1 (low) | 365 / 1794 | 1.99 (1.66, 2.40) | 2.01 (1.67, 2.43) | 1.99 (1.66, 2.40) | 2.01 (1.67, 2.42) | 1.64 (1.35, 1.99) | 1.67 (1.37, 2.03) |
| P for trend |  | p < 0.0001 | p < 0.0001 | p < 0.0001 | p < 0.0001 | p < 0.0001 | p < 0.0001 |
| Per SD decrease | 953 / 7361 | 1.76 (1.62, 1.90) | 1.77 (1.63, 1.91) | 1.76 (1.62, 1.90) | 1.78 (1.64, 1.91) | 1.52 (1.37, 1.67) | 1.54 (1.40, 1.69) |

Numbers of study members in this sample corresponds to those with complete data on all variables in the analyses. Thresholds for categories of cognitive function: tertile 1 ( $\geq 108.3$ ); tertile 2 (108.2-93.3); and tertile 1 ( $\geq 93.2$ ). A standard deviation (SD) in general cognitive function was 15 units.

To gain some information about whether the association with vaccine hesitancy was linear, the analyses were repeated according to tertiles of cognitive function. In age-, sex- and ethnicity-adjusted analyses, relative to people in the highest-scoring cognition tertile, those in the lowest were twice as likely to be vaccine hesitant (1.99; 1.66, 2.40). People in the intermediate ability tertial had intermediate risk of hesitancy (1.28; 1.07, 1.54) such that an incremental effect was apparent across the cognition categories (p-value for trend: <0.0001). Lastly, in order to explore inflections in the cognition–hesitancy association, we utilised deciles of cognition in analyses. Again, there was evidence of a clear trend, although this was not perfectly stepwise across all categories (figure 2).

**Figure 2.**
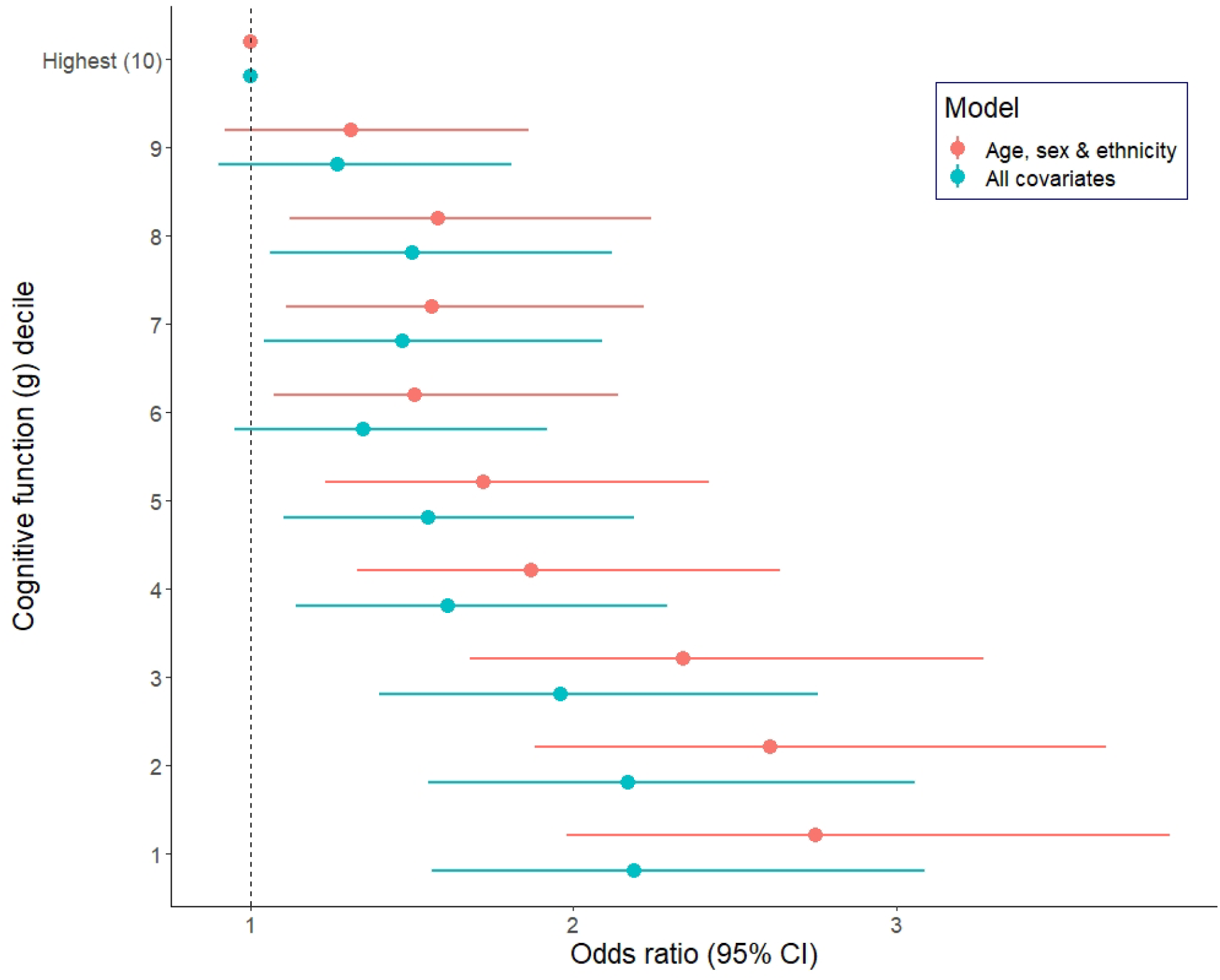
Odds ratios (95% CI) for the relation of general cognitive function with COVID-19 vaccine hesitancy in Understanding Society. All covariates are: age, sex, ethnicity, somatic comorbidity, psychological distress, shielding, and education. In both models, the p-value for trend across deciles was <0.0001

## Discussion

Our main finding was that, in data collected immediately following the announcement in the UK of an efficacious vaccine, and net of several covariates, people with lower scores on tests of cognitive function were less minded to take up an offer of vaccination for COVID-19 if it was made. These effects were apparent across the full range of cognition scores. That we were able to replicate known predictors of COVID-19 vaccine hesitancy – being female,^36-38^ younger,^36,38^ non-white ethnicity,^21,38,39^ and having a lower prevalence of physical morbidity^40^ – gives us some confidence in our novel results for cognitive function.

### Comparison with existing studies

To the best of our knowledge, there has been one prior examination of the relationship between cognition and vaccine hestinacy.^16^ Comprising two small cross-sectional studies where data collection took place before the announcement of vaccine test results, people who performed better on a short test of cognitive function were more likely to be vaccine-accepting.^16^ We found similar results in using more detailed measures of cognitive function in a large sample which allowed us to explore the shape of the relationship across the full range of abilities. A closely related literature is that for education with which cognition is positively correlated.^41^ This supports the present results whereby people with higher educational achievement were less likely to be vaccine-hesitant. ^42,43^

### Potential public health implications

We have recently shown that, in data collected prior to vaccine distribution, of a range of baseline psychosocial factors which included socioeconomic status, education, personality type, and mental health, cognitive function was the most strongly associated with subsequent incidence of severe COVID-19, such that a doubling of the risk of hospitalisation was apparent in the lowest scoring group.^44^ This supports other data that individuals with higher cognitive function experience a lower risk of death from other respiratory diseases, including influenza and pneumonia.^45^ The notion that people with lower cognitive ability appear to have greater rates of severe COVID-19^44,46^ and, based on the present results are also less likely to take up the offer of vaccination, may further increase the disease burden in this group, as may also be the case for people from ethnic minority groups^21,47^ and the socioeconomically disadvantaged.^16,44^

### Plausible explanations

Various explanations may be germane to the cognition–vaccine intention link, including the observation that people with higher cognitive ability are better equipped to obtain, process, and respond to disease prevention advice.^48^ There has been a deluge of health advice in the current pandemic during an era when news outlets and social media platforms have never been more ubiquitous and influential. Preventative information has ranged from the simple and practical to the complex, contradictory, false, and fraudulent. In order to diminish their risk of the infection, people have to acquire, synthesise, weigh-up, and deploy this information but the ability to do so seems to vary by levels of health literacy^49^ just as it may for its close correlate, cognitive function.

### Study strengths and weaknesses

While the present study has its strengths, including its size, national representativeness, and timing, there are also some weaknesses. First, we used vaccine intentionality as an indicator vaccine uptake but the correlation is imperfect. In a small scale longitudinal study conducted during the period of the 2009 H1N1 pandemic in Hong Kong, less than 10% of people who expressed a commitment to being inoculated reported that they had received a vaccination two months later.^50^ Elsewhere, in a US adult population at high risk of seasonal influenza, around half of those intending to be vaccinated had received the inoculation within the following 5 months.^51^ Second, there was inevitably some loss to follow-up (figure 1). While this attrition may have impacted upon the estimation of the prevalence vaccine hesitancy which is likely to be lower in our select sample relative to the general population,^52^ it is unlikely to have influenced our estimation of its relationship with cognitive function. Thus, in other contexts, we have shown that highly select cohorts reveal very similar risk factor–disease associations to those seen in studies with conventionally high response.^53^

In conclusion, people with lower scores on standard tests of cognitive function reported being less willing to take up the future offer of vaccination for COVID-19. It is possible that erroneous social media news reports have complicated decision-making. Special efforts should be made to communicate clear information about vaccine efficacy and safety so that everyone—including those who report being less likely to choose vaccination—can make well-informed choices.

## Data Availability

Researchers who would like to use Understanding Society data need to register with the UK Data Service (https://www.ukdataservice.ac.uk/) before application.

## Contributions

GDB generated the idea for the present manuscript. DA built the dataset, conducted all analyses, and prepared the displayable items. All authors developed an analytical plan and commented on a manuscript drafted by GDB.

## References

1. Deary IJ, Batty GD. Cognitive epidemiology. J Epidemiol Community Health 2007;61(5):378–384.

2. Deary IJ. Intelligence. Annu Rev Psychol 2012;63:453–82.

3. Gottfredson LS. Intelligence: is it the epidemiologists’ elusive “fundamental cause” of social class inequalities in health? J. Pers. Soc. Psychol 2004;86(1):174–199.

4. Batty GD, Deary IJ, Schoon I, Gale CR. Childhood mental ability in relation to food intake and physical activity in adulthood: the 1970 British Cohort Study. Pediatrics 2007;119(1):e38–e45.

5. Whalley L, Fox H, Lemmon H, Duthie S, Collins A, Peace H, Starr J, Deary I. Dietary supplement use in old age: associations with childhood IQ, current cognition and health. International journal of geriatric psychiatry 2003;18(9):769–776.

6. Batty GD, Deary IJ, Schoon I, Gale CR. Mental ability across childhood in relation to risk factors for premature mortality in adult life: the 1970 British Cohort Study. J Epidemiol Community Health 2007;61(11):997–1003.

7. Batty GD, Deary IJ, MacIntyre S. Childhood IQ in relation to risk factors for premature mortality in middle-aged persons: the Aberdeen Children of the 1950s study. J Epidemiol Community Health 2007;61(3):241–247.

8. Batty GD, Deary IJ, MacIntyre S. Childhood IQ and life course socioeconomic position in relation to alcohol induced hangovers in adulthood: the Aberdeen children of the 1950s study. J Epidemiol Community Health 2006;60(10):872–874.

9. Batty GD, Deary IJ, Schoon I, Emslie C, Hunt K, Gale CR. Childhood mental ability and adult alcohol intake and alcohol problems: the 1970 British cohort study. Am J Public Health 2008;98(12):2237–2243.

10. Taylor MD, Hart CL, Smith GD, Starr JM, Hole DJ, Whalley LJ, Wilson V, Deary IJ. Childhood IQ and social factors on smoking behaviour, lung function and smoking-related outcomes in adulthood: linking the Scottish Mental Survey 1932 and the Midspan studies. Br J Health Psychol 2005;10(Pt 3):399–410.

11. Deary IJ, Gale CR, Stewart MC, Fowkes FG, Murray GD, Batty GD, Price JF. Intelligence and persisting with medication for two years: Analysis in a randomised controlled trial. Intelligence 2009;37(6):607–612.

12. Wallert J, Lissåker C, Madison G, Held C, Olsson E. Young adulthood cognitive ability predicts statin adherence in middle-aged men after first myocardial infarction: A Swedish National Registry study. Eur J Prev Cardiol 2017;24(6):639–646.

13. O’Conor R, Muellers K, Arvanitis M, Vicencio DP, Wolf MS, Wisnivesky JP, Federman AD. Effects of health literacy and cognitive abilities on COPD self-management behaviors: A prospective cohort study. Respir Med 2019;160:105630.

14. Gale CR, Deary IJ, Wardle J, Zaninotto P, Batty GD. Cognitive ability and personality as predictors of participation in a national colorectal cancer screening programme: the English Longitudinal Study of Ageing. J Epidemiol Community Health 2015;69(6):530–535.

15. Omer SB, Yildirim I, Forman HP. Herd immunity and implications for SARS-CoV-2 control. Jama 2020;324(20):2095–2096.

16. Murphy J, Vallières F, Bentall RP, Shevlin M, McBride O, Hartman TK, McKay R, Bennett K, Mason L, Gibson-Miller J, Levita L, Martinez AP, Stocks TVA, Karatzias T, Hyland P. Psychological characteristics associated with COVID-19 vaccine hesitancy and resistance in Ireland and the United Kingdom. Nat Commun 2021;12(1):29.

17. Gallacher J. Covid-19: Oxford University vaccine is highly effective (BBC News). https://www.bbc.co.uk/news/health-55040635 2020.

18. YouGov ICL. COVID-19 Behaviour Tracker. https://ichpanalytics.imperialcollegehealthpartners.com/t/BDAU/views/YouGovICLCOVID-19BehaviourTracker/4Allbehaviorsovertime?:iid=1&:isGuestRedirectFromVizportal=y&:embed=y 2021.

19. Lynn P. Sample design for understanding society. Underst. Soc. Work. Pap. Ser 2009;2009.

20. Burton J, Lynn P, Benzeval M. How Understanding Society: The UK household longitudinal study adapted to the COVID-19 pandemic. Survey Research Methods, 2020;235–239.

21. Robertson E, Reeve KS, Niedzwiedz CL, Moore J, Blake M, Green M, Katikireddi SV, Benzeval MJ. Predictors of COVID-19 vaccine hesitancy in the UK Household Longitudinal Study. Brain Behavior Immunity 2021:2020.12. 27.20248899.

22. McFall S. Understanding Society: UK household longitudinal study: Cognitive ability measures. Institute for Social and Economic Research, University of Essex 2013.

23. Gray M, D’Ardenne J, Balarajan M, Uhrig N. Cognitive testing of wave 3 understanding society questions. Institute for Social and Economic Research: University of Essex 2011.

24. Steptoe A, Breeze E, Banks J, Nazroo J. Cohort Profile: The English Longitudinal Study of Ageing. Int. J. Epidemiol 2013;42(6):1640–1648.

25. Borsch-Supan A, Brandt M, Hunkler C, Kneip T, Korbmacher J, Malter F, Schaan B, Stuck S, Zuber S. Data Resource Profile: the Survey of Health, Ageing and Retirement in Europe (SHARE). Int. J. Epidemiol 2013;42(4):992–1001.

26. Richards M, Shipley B, Fuhrer R, Wadsworth ME. Cognitive ability in childhood and cognitive decline in mid-life: longitudinal birth cohort study. Bmj 2004;328(7439):552.

27. Sonnega A, Faul JD, Ofstedal MB, Langa KM, Phillips JW, Weir DR. Cohort Profile: the Health and Retirement Study (HRS). Int. J Epidemiol 2014;43(2):576–585.

28. Lachman ME, Agrigoroaei S, Murphy C, Tun PA. Frequent cognitive activity compensates for education differences in episodic memory. The American Journal of Geriatric Psychiatry 2010;18(1):4–10.

29. Crum RM, Anthony JC, Bassett SS, Folstein MF. Population-based norms for the Mini-Mental State Examination by age and educational level. JAMA 1993;269(18):2386–2391.

30. Huppert FA, Brayne C, Gill C, Paykel E, Beardsall L. CAMCOG—A concise neuropsychological test to assist dementia diagnosis: Socio-demographic determinants in an elderly population sample. British Journal of Clinical Psychology 1995;34(4):529–541.

31. Holi MM, Marttunen M, Aalberg V. Comparison of the GHQ-36, the GHQ-12 and the SCL-90 as psychiatric screening instruments in the Finnish population. Nord. J. Psychiatry 2003;57(3):233–238.

32. Hankins M. The factor structure of the twelve item General Health Questionnaire (GHQ-12): the result of negative phrasing? Clinical Practice and Epidemiology in Mental Health 2008;4(1):1–8.

33. Russ TC, Stamatakis E, Hamer M, Starr JM, Kivimaki M, Batty GD. Association between psychological distress and mortality: individual participant pooled analysis of 10 prospective cohort studies. BMJ 2012;345:e4933.

34. Russ T, Hamer M, Stamatakis E, Starr J, Batty G. Psychological Distress as a Risk Factor for Dementia Death. Arch Intern Med 2011;171:1858–1859.

35. Russ TC, Kivimaki M, Morling JR, Starr JM, Stamatakis E, Batty GD. Association Between Psychological Distress and Liver Disease Mortality: A Meta-analysis of Individual Study Participants. Gastroenterology 2015;148(5):958–966.

36. Detoc M, Bruel S, Frappe P, Tardy B, Botelho-Nevers E, Gagneux-Brunon A. Intention to participate in a COVID-19 vaccine clinical trial and to get vaccinated against COVID-19 in France during the pandemic. Vaccine 2020;38(45):7002–7006.

37. Wang J, Jing R, Lai X, Zhang H, Lyu Y, Knoll MD, Fang H. Acceptance of COVID-19 Vaccination during the COVID-19 Pandemic in China. Vaccines (Basel) 2020;8(3).

38. Freeman D, Loe BS, Chadwick A, Vaccari C, Waite F, Rosebrock L, Jenner L, Petit A, Lewandowsky S, Vanderslott S, Innocenti S, Larkin M, Giubilini A, Yu LM, McShane H, Pollard AJ, Lambe S. COVID-19 vaccine hesitancy in the UK: the Oxford coronavirus explanations, attitudes, and narratives survey (Oceans) II. Psychol Med 2020:1–15.

39. Williams L, Flowers P, McLeod J, Young D, Rollins L. Social patterning and stability of intention to accept a COVID-19 vaccine in Scotland: Will those most at risk accept a vaccine? Vaccines 2021;9(1):17.

40. Ruiz JB, Bell RA. Predictors of intention to vaccinate against COVID-19: Results of a nationwide survey. Vaccine 2021.

41. Neisser U, Boodoo G, Bouchard Jnr T, Boykin AW, Brody N, Ceci SJ, Halpern DF, Loehlin JC, Perloff R, Sternberg RJ, Urbina S. Intelligence: knowns and unknowns. Am Psychol 1996;51:77–101.

42. Kuter BJ, Browne S, Momplaisir FM, Feemster KA, Shen AK, Green-McKenzie J, Faig W, Offit PA. Perspectives on the receipt of a COVID-19 vaccine: A survey of employees in two large hospitals in Philadelphia. Vaccine 2021.

43. Nguyen KH, Srivastav A, Razzaghi H, Williams W, Lindley MC, Jorgensen C, Abad N, Singleton JA. COVID-19 Vaccination Intent, Perceptions, and Reasons for Not Vaccinating Among Groups Prioritized for Early Vaccination - United States, September and December 2020. MMWR Morb Mortal Wkly Rep 2021;70(6):217–222.

44. Batty GD, Deary IJ, Luciano M, Altschul DM, Kivimaki M, Gale CR. Psychosocial factors and hospitalisations for COVID-19: Prospective cohort study based on a community sample. Brain Behav Immun 2020;89:569–578.

45. Gale CR, Deary IJ, Batty GD. Cognitive ability and risk of death from lower respiratory tract infection: findings from UK Biobank. Sci Rep 2019;9(1):1342.

46. Batty GD, Deary I, Gale C. Pre-pandemic cognitive function and COVID-19 mortality: prospective cohort study. medRxiv 2021.

47. Lassale C, Gaye B, Hamer M, Gale CR, Batty GD. Ethnic disparities in hospitalisation for COVID-19 in England: The role of socioeconomic factors, mental health, and inflammatory and pro-inflammatory factors in a community-based cohort study. Brain Behav Immun 2020;88:44–49.

48. von Wagner C, Steptoe A, Wolf MS, Wardle J. Health literacy and health actions: a review and a framework from health psychology. Health Educ Behav 2009;36(5):860–77.

49. Wolf MS, Serper M, Opsasnick L, O’Conor RM, Curtis LM, Benavente JY, Wismer G, Batio S, Eifler M, Zheng P, Russell A, Arvanitis M, Ladner D, Kwasny M, Persell SD, Rowe T, Linder JA, Bailey SC. Awareness, Attitudes, and Actions Related to COVID-19 Among Adults With Chronic Conditions at the Onset of the U.S. Outbreak: A Cross-sectional Survey. Ann Intern Med 2020.

50. Liao Q, Cowling BJ, Lam WW, Fielding R. Factors affecting intention to receive and self-reported receipt of 2009 pandemic (H1N1) vaccine in Hong Kong: a longitudinal study. PLoS One 2011;6(3):e17713.

51. Harris KM, Maurer J, Lurie N. Do people who intend to get a flu shot actually get one? J Gen Intern Med 2009;24(12):1311–3.

52. Fry A, Littlejohns TJ, Sudlow C, Doherty N, Adamska L, Sprosen T, Collins R, Allen NE. Comparison of Sociodemographic and Health-Related Characteristics of UK Biobank Participants With Those of the General Population. Am J Epidemiol 2017;186(9):1026–1034.

53. Batty GD, Gale CR, Kivimaki M, Deary IJ, Bell S. Comparison of risk factor associations in UK Biobank against representative, general population based studies with conventional response rates: prospective cohort study and individual participant meta-analysis. BMJ 2020;368:m131.

